# A randomized, double□blind, placebo□controlled study evaluating the impact of *Hericium erinaceus* (Lion’s Mane) on cognitive performance and subjective wellbeing

**DOI:** 10.64898/2026.04.13.26350781

**Authors:** J. Daoust, S. Farrar, A.D. Grant, M.C.B. Erfe, P.L. Oliver, V. Luna, J. Moos, N. Craft

**Affiliations:** M2 Ingredients, Vista, CA, USA; People Science, Inc., Los Angeles, CA, USA

**Keywords:** Cognition, attention, sleep, mood, functional foods, working memory

## Abstract

*Hericium erinaceus* (Lion’s Mane) is a functional mushroom with a long history of culinary and traditional use, as well as potential neurotrophic and mood□modulating properties. Evidence for its effects on cognitive performance under real□world conditions, however, remains limited. In this randomized, double□blind, placebo□controlled trial, adults aged 40–75 years with self□reported cognitive difficulty completed a one□week baseline followed by eight weeks of daily supplementation with 2 g of *H. erinaceus* fruiting body and mycelial biomass or placebo. Cognitive performance using a computerized battery, as well as daily subjective assessments of sleep and wellbeing, were collected remotely. 109 Participants were included in the primary analysis (*H. erinaceus*, n = 57; placebo, n = 52). *H. erinaceus* was associated with significantly greater improvement in visual attention and working memory (Juggle Factor task), subjective sleep quality, morning restedness, and mood compared with placebo (*p* < 0.05). No adverse events were reported in participants receiving *H. erinaceus*. Together, *H. erinaceus* supplementation modestly improved visual attention and was associated with faster improvements in sleep quality, restedness, and mood in adults with subjective cognitive concerns.

## 1. Introduction

*Hericium erinaceus* (Lion’s Mane) is an edible basidiomycete mushroom widely consumed as both a food and dietary supplement. It has a long history of traditional use in East Asia as a medicinal□culinary mushroom, where it has been valued for supporting digestion, vitality, and cognitive health (Qi, 2024). In recent years, scientific interest has increasingly focused on its potential neurotrophic, neuroprotective, and psychotropic properties (Cipriano et al., 2026; Contato & Conte-Junior, 2025; Mori et al., 2008).

A diverse array of bioactive compounds has been identified in *H. erinaceus*, including erinacines, hericenones, and related meroterpenoids, which differ in abundance between fruiting body and mycelial preparations (Qiu et al., 2024). Preclinical studies have demonstrated that these compounds can stimulate nerve growth factor (NGF) and brain□derived neurotrophic factor (BDNF) synthesis, promote neurite outgrowth, and activate intracellular signaling pathways relevant to neuroplasticity, including ERK1/2 signaling (Martínez-Mármol et al., 2023; Qiu et al., 2024). These mechanisms provide a biological rationale for preliminary effects on cognition, mood, and sleep.

In animal models, supplementation with *H. erinaceus* has been shown to improve recognition and to induce hippocampal and cerebellar neurogenesis during aging (Ratto et al., 2019). Additional preclinical evidence suggests that *H. erinaceus* may exert anti□inflammatory and neuroprotective effects in preclinical disease models, further supporting its potential relevance to brain health (Chau et al., 2023).

Clinical investigations of *H. erinaceus* supplementation have reported improvements in mood, anxiety, sleep quality, and mixed results across domains of cognitive performance, with outcomes varying across populations, study durations, and ingredient formats (Černelič Bizjak et al., 2024, 2024; Docherty et al., 2023; Nagano et al., 2010; Saitsu et al., 2019; Spangenberg et al., 2025; Vigna et al., 2019). Differences in cultivation methods and the use of fruiting body versus mycelium may contribute to variability in bioactive profiles and study findings. Preparations incorporating both fruiting body and mycelial biomass may therefore provide broader exposure to bioactives produced across the fungal life cycle.

Beyond direct neurotrophic effects, emerging evidence suggests that *H. erinaceus* may also influence cognitive and emotional outcomes indirectly through modulation of the gut microbiota and production of neuroactive metabolites (Daoust et al., 2025). Such gut–brain axis mechanisms may be particularly relevant for functional food applications.

Decentralized clinical trial methods provide an opportunity to evaluate functional food ingredients under real□world conditions, improving external validity while enabling frequent longitudinal assessment. Accordingly, the present randomized, double□blind, placebo□controlled study evaluated the effects of daily supplementation with 2 g of *H. erinaceus* fruiting body and mycelial biomass for eight weeks on cognitive performance and subjective wellbeing in adults aged 40–75 years with self□reported cognitive difficulty.

### 2. Methods

### 2.1 Eligibility

Participants were included if they met the following inclusion criteria: i) aged 40-75 years old, ii) self-reported concerns of reduced memory, focus and cognition, iii) a Cognitive Failures Questionnaire score of mild-to-moderate at screening, iv) interest in use of the study product and in tracking their own cognition, v) willingness to do a 4-week washout from any memory or cognitive-function-targeted supplements prior to randomization, vi) willingness to do a 4 week washout from any mushroom supplement compound, vii) if taking any medications for sleep in the 4 weeks prior to enrollment and during the study, willingness to maintain a stable dose, viii) willing to avoid large deviations in alcohol consumption and to log alcohol consumption during the study.

Participants were also required to meet several lifestyle and behavioural inclusion criteria, including general good health at screening (Investigator discretion), English fluency, ability to provide informed consent, use a personal smartphone and computer, receive the study product and wearable device at an address in the US, complete study assessments over the course of up to 9 weeks, wear and sync an Oura ring sleep monitoring device daily.

Participants were excluded if they were receiving cognitive behavioral therapy for insomnia (CBTi), any investigational therapies in the 30 days prior to randomization, or had any of the following: Alzheimer’s or dementia, a neurodevelopmental disorder (e.g., dyslexia or ADHD), visual or auditory impairment, narcolepsy, restless leg syndrome, circadian rhythm disorders, psychotic disorder, substance abuse disorder, pregnancy or planned pregnancy, long Covid, allergy to functional mushrooms, or were users of nicotine or cannabis. Final exclusion was at the investigator’s discretion.

### 2.2 Study Design

Participants completed a 14-week study consisting of screening period, randomization and shipping period, a 1-week baseline period, and an 8-week Lion’s Man/placebo use period (**See Table 1**). Participants were randomized to one of two groups: A or B: (A) Lion’s mane (2g/ 3 capsules daily), and (B) matching placebo capsules. The Investigators, study team and participants were blinded to group assignment. Participants received the study product or placebo and study supplies after randomization. Demographic and medical history data, including concomitant medications, were collected during the screening period. Survey data, as described below, were collected during baseline and the probiotic study product/placebo use period. Additionally, participants completed a suite of cognitive tasks using the online platform BrainHQ (Posit Science Corp, San Francisco, CA, USA). All subjective data were collected through a mobile research app, the Consumer Health Learning & Organizing Ecosystem (Chloe) (People Science, Inc., Los Angeles, CA, USA), described below.

**Table 1.** Demographics.

| **Category** | **Total % (Count)** |
| --- | --- |
| **Sex** |  |
| Female | 52.3 (57) |
| Male | 47.7 (52) |
| **Mean Age (STD)** | 53.5 (9.1) |
| **Ethnicity** |  |
| Caucasian | 68.8 (75) |
| African American | 12.8 (14) |
| Asian | 9.17 (10) |
| Hispanic | 6.42 (7) |
| Native American or<br>Alaska Native | 0.917 (1) |
| Other | 1.83 (2) |

### 2.3 Data Management System

All data was securely stored on Chloe Amazon Web Services HIPAA compliant servers. The Chloe platform contains modules for building and managing surveys, study landing pages, marketing outreach with tracking tools for recruitment, audited electronic consent forms, data management and analytics using an integrated relational database. Additionally, the platform contains a user-facing app for participants that delivers surveys, study instructions, calendar reminders, communication with study team, and personal data reports at study culmination. Data from completed assessments was automatically collected for analysis.

### 2.4 Recruitment, Consent, and Enrollment

Participants were recruited through social media channels and researcher networks. Recruitment outreach consisted of IRB-approved advertising by email, digital marketing channels and word of mouth. The IRB-approved study landing page hosted on the People Science website led to an IRB-approved pre-screening questionnaire to determine individual qualification. Qualified participants were invited to download the Chloe application (described below) and create an account to access the Informed Consent Form and continue through eligibility determination and enrollment.

Virtual electronic informed consent, including a study specific privacy authorization and the California Experimental Subject’s Bill of Rights (as applicable) were provided through the HIPAA-compliant cloud-based platform Chloe. Eligible participants who provided virtual electronic consent were automatically registered into the study by the platform.

### 2.5 Intervention

The test product was provided by M2 Ingredients Inc. (Vista, California, USA) and consisted of certified organic powdered *H. erinaceus* preparation containing both fruiting body and mycelial biomass (strain M2□102□10), produced via solid□state fermentation cultured on whole certified organic oats (*Avena sativa*) (Alemayehu et al., 2023), dehydrated and then milled into a powder. The finished powders contained <6 % moisture and had a particle size of ≥95 % through a 60-mesh screen (250 μm). Identities were confirmed with species positive identification specifications using DNA sequencing of the master tissue culture, taxonomic and visual monitoring of morphology, and growth metrics during the growing cycle. The powder was encapsulated in vegetable cellulose capsules. Participants consumed 2 g daily (three capsules). Placebo capsules were visually matched and filled with 2g of acacia gum.

### 2.6 Study Timeline and Activities

Participants’ activities were as follows **(See Supplemental Table 1)**. Participants who gave digital Informed Consent continued on to the Screening period where they completed demographics, medical history, concomitant medications and the Cognitive Failures Questionnaire (CFQ). Eligible participants were randomized to Lion’s Mane (2 g powder /3 capsule dose, see section 2.5 Intervention, above) or matching placebo and were shipped their study supplies. All randomized participants confirmed shipment receipt before proceeding to the baseline period. Participants completed a 1-week baseline, followed by 8 weeks of Lion’s Mane or placebo use. Participants were prompted to report any adverse events (AEs) weekly or advised to reach out to the study team at any other time to report an AE.

### 2.7 Cognitive Tasks

BrainHQ (Posit Science Corp, San Francisco, CA) an online cognitive task administration tool, was used to evaluate participant cognitive performance across the study. The BrainHQ suite was administered at biweekly intervals (**See Supplemental Table 1**) on a participant’s home computer. Descriptions of tasks are presented in **Supplemental Table 2**, and were analyzed individually as well as via a composite battery score. Participants were instructed to complete the assessments in a distraction-free environment. Task performance was analyzed both using values that were normed (z-score) to the entire database of participants who had tested this BrainHQ suite, as well as raw units corresponding to each test as follows: Juggle Factor (number of objects), Double Decision (ms), Target Tracker (number of objects), Memory Grid (number of matched cards), Mental Map (map complexity level), Composite Suite of the preceding listed tasks (percentile).

### 2.8 Subjective Daily Assessments

Participants completed daily numeric rating scales (1-10) of sleep quality, morning restedness, overall mood, stress, motivation and focus. Participants also reported daily adherence to product use, any changes to daily medications, as well as consumption of alcohol or evening caffeine.

### 2.9 Adverse Events

Participants were prompted weekly to disclose any adverse events that occurred.

### 2.10 Data Evaluability, Analysis and Statistics

Participants were required to fill out their baseline and study product/placebo use period cognitive tasks. Individual days of data were excluded if the product/placebo consumption was missed in analysis of daily surveys. These exclusions were made prior to unblinding. All analyses were conducted using Python Jupyter Notebooks (Project Jupyter, Beaverton, OR, USA). Normality was assessed using the Shapiro-Wilk (SW) test. Non-parametric statistics were used to compare specific timepoints as data were not normally distributed (i.e., for analysis of BrainHQ data and daily data). BrainHQ data were analyzed both in their raw format, as well as in within-individual and Brain-HQ user base-wide, population-normed formats. All cognitive test data were assessed for baseline differences, and adjusted for normative headroom as appropriate (**See: Supplemental Methods**). Benjamini-Hoch (BH) correction was used in the case of multiple comparison testing. Linear mixed models (LMM) were used to evaluate potential impacts of demographic factors on outcomes. Mann-Kendall (MK) tests were used to evaluate potential trends over time in daily data. Fisher’s Exact Tests (FET) were used to evaluate differences in adverse event occurrences. Reaction time variability was not available for analysis due to differences in participant browser and device settings.

## 3. Results

### 3.1 Recruitment and Conduct

This study was approved by Sterling Institutional Review Board (Sterling IRB ID 13098) and registered with ClinicalTrials.gov (NCT06870136). All participants gave informed consent.

### 3.2 Sample Size and Demographics

Of 150 participants who were randomized, 120 completed the study and 30 were withdrawn due to loss to follow-up (21), participant decision (8), and investigator’s decision (1). N=109 participants were evaluated (Lion’s Mane=57, Placebo=52). Demographics are described in **Table 1**.

### 3.3 Cognitive Performance Primary Outcome

Participants receiving *H. erinaceus* demonstrated significantly greater improvement over time on the Juggle Factor task compared with placebo (linear mixed model, *p* = 0.001, +0.45 z scores). No significant between□group differences were observed for Memory Grid, Target Tracker, Mental Map, Sandia’s Matrices, or composite cognitive scores. An apparent between-group difference favoring placebo was observed in the Double Decision reaction time task in unadjusted analyses at Week 8. However, this effect was not statistical after accounting for baseline performance differences and ceiling effects, suggesting it was primarily driven by greater headroom for improvement in the placebo group (data not shown).

### 3.4 Subjective Sleep Quality, Restedness and Mood

Participants in the *H. erinaceus* group exhibited a faster rate of improvement in sleep quality, morning restedness, and mood compared with placebo (*p* < 0.05). Daily measures did not differ in any metric at baseline. The placebo group improved at a faster rate with respect to subjective motivation (LMM p<0.05).

### 3.5 Safety

No adverse events were reported in the *H. erinaceus* group. Three mild□to□moderate adverse events occurred in the placebo group (Table 2).

**Table 2.** Adverse Events.

| **Group** | **Number**<br>**of AEs** | **Description** | **Severity** | **Note** |
| --- | --- | --- | --- | --- |
| **Placebo** | 2 | Dizziness | Both mild | Both resolved |
| **Placebo** | 1 | Headache | Moderate | Resolved |

## 4. Discussion

This randomized, double□blind, placebo□controlled study demonstrates that daily supplementation with *H. erinaceus* fruiting body and mycelial biomass may modestly improve visual attention and working memory, as well as rate of subjective sleep quality, restedness, and mood compared to placebo. Previous cognitive benefit reports are mixed, potentially due to smaller sample sizes, variable task timing after consumption, and variation in dose and portion of the Lion’s Mane mushroom consumed. (Cha et al., 2024). The present findings are consistent with prior clinical literature reporting mood and sleep benefits of *H. erinaceus* supplementation, as well as with reports of working memory and visual attention benefits (Černelič Bizjak et al., 2024; Docherty et al., 2023; Nagano et al., 2010; Saitsu et al., 2019; Surendran et al., 2025; Vigna et al., 2019).

The observed effects are biologically plausible in light of preclinical evidence demonstrating neurotrophic, neurogenic, and anti□inflammatory mechanisms associated with *H. erinaceus* bioactives (Chau et al., 2023; Martínez-Mármol et al., 2023; Qiu et al., 2024; Ratto et al., 2019). In addition, emerging data suggest that modulation of the gut microbiota may contribute to downstream cognitive and emotional outcomes, highlighting a potential gut– brain axis component(Daoust et al., 2025) .

The decentralized study design enabled assessment of both acute and chronic impacts of Lion’s mane on a larger sample under real□world conditions, enhancing relevance for functional food applications. Limitations to the study design include the potential for increased variability relative to site□based trials. Importantly, no product□related adverse events were observed, supporting the safety and tolerability of the intervention.

## 5. Conclusions

Supplementation with *Hericium erinaceus* fruiting body and mycelial biomass for eight weeks was safe and was associated with modest, clinically significant improvements in visual attention and subjective wellbeing in adults with self□reported cognitive concerns. These findings support further investigation of *H. erinaceus* as a functional food ingredient for cognitive and emotional health.

**Figure 1.**
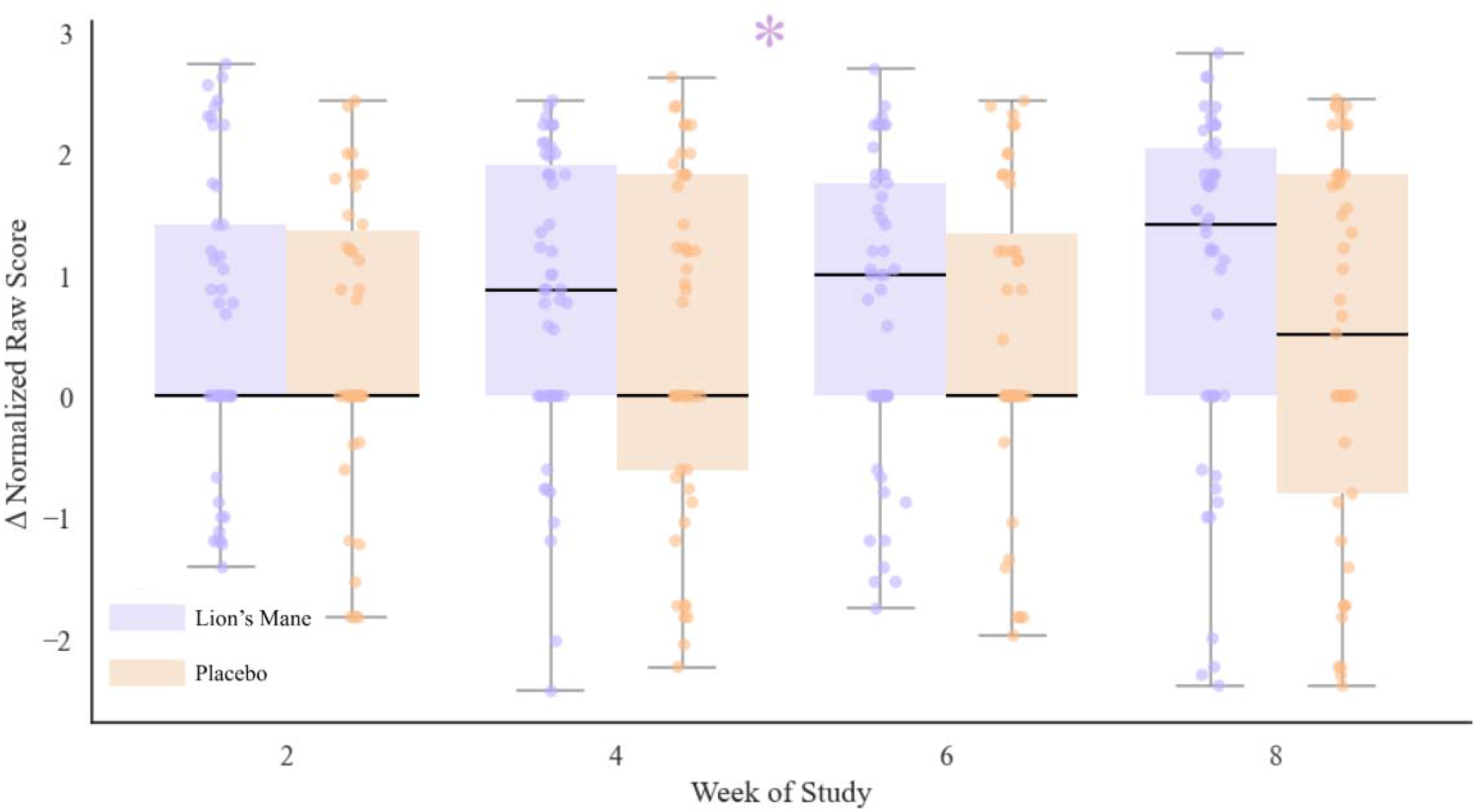
Juggle Factor Improved to a Greater Degree with Lion’s Mane Consumption Across the Study Period. Box and whiskers of within-individual change by week between groups in normalized Juggle Factor Scores. Individual dots represent individual data. Lion’s Mane is shown in purple and Placebo is shown in orange. Overall improvement across the study was statistically greater in Lion’s Mane (LMM p=0.001). No individual time point differed statistically.

**Figure 2.**
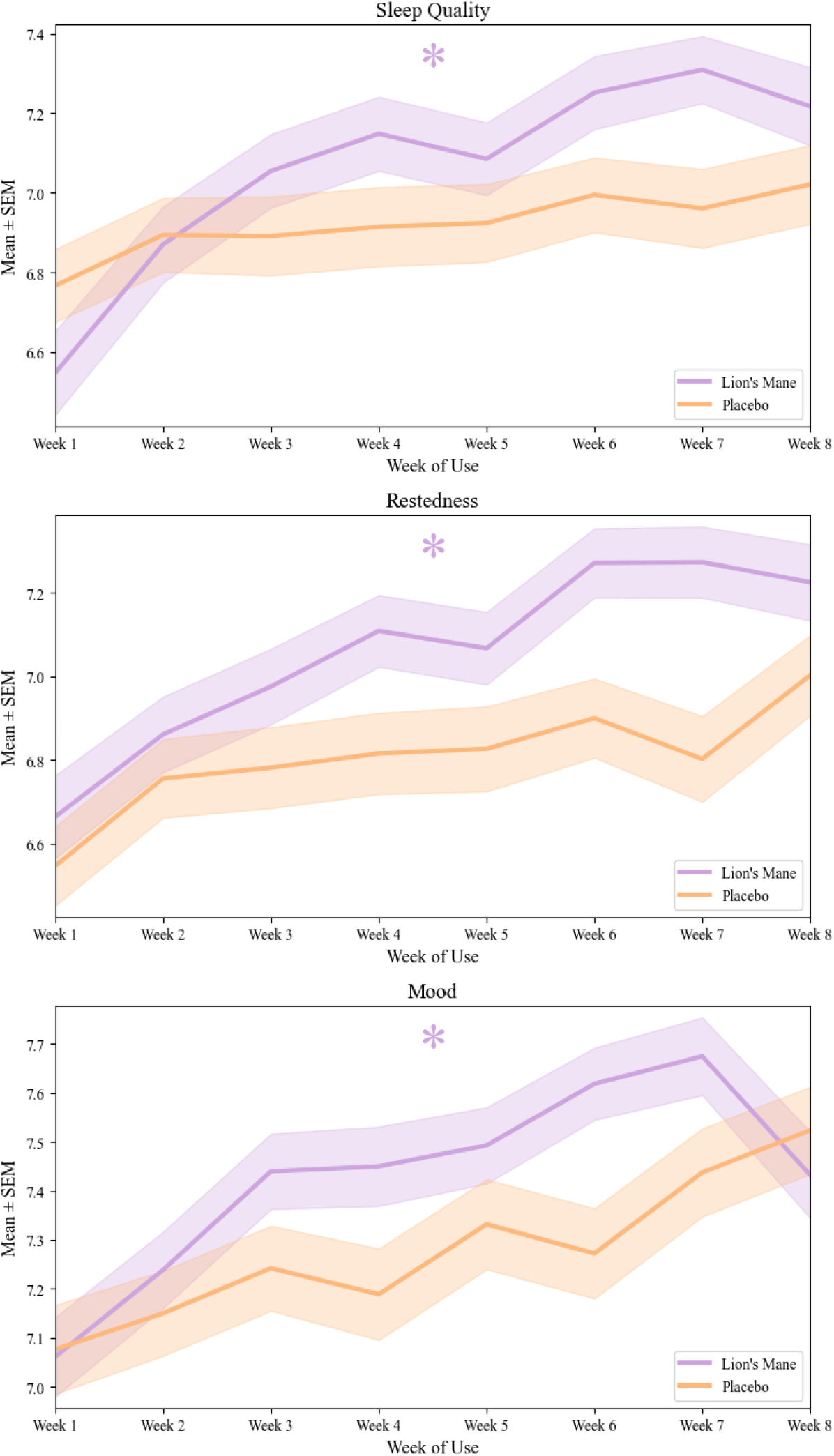
Lion’s Mane Daily Data Across the Study Reveals Steeper Rate of Improvement in Sleep Quality, Restedness and Mood. Daily subjective data reveals a faster rate of improvement in Lion’s Mane for Sleep Quality (top), Restedness (middle) and Mood (bottom). (LMM of slope per day by group in weekly averaged data p<0.05 in each case).

## Data Availability

Data are available upon reasonable request from the corresponding author.

## Acknowledgements

The authors would like to thank Dr. Caitlin Stamatis for her editorial input on this manuscript.

